# Forecasting drug overdose mortality by age in the United States at the national and county levels

**DOI:** 10.1101/2023.09.25.23296097

**Authors:** Lucas Böttcher, Tom Chou, Maria R. D’Orsogna

## Abstract

The drug overdose crisis in the United States continues to intensify. Fatalities have increased five-fold since 1999 reaching a record high of 108,000 deaths in 2021. The epidemic has unfolded through distinct waves of different drug types, uniquely impacting various age, gender, race and ethnic groups in specific geographical areas. One major challenge in designing effective interventions is the forecasting of age-specific overdose patterns at the local level so that prevention and preparedness can be effectively delivered. We develop a forecasting method that assimilates observational data obtained from the CDC WONDER database with an age-structured model of addiction and overdose mortality. We apply our method nationwide and to three select areas: Los Angeles County, Cook County and the five boroughs of New York City, providing forecasts of drug-overdose mortality and estimates of relevant epidemiological quantities, such as mortality and age-specific addiction rates.

**Significance:** The drug overdose epidemic in the United States continues to escalate, with fatalities increasing five-fold since 1999 and reaching a record high of 108,000 individuals in 2021. The crisis is characterized by distinct waves of drug types, disproportionately affecting various demographic groups in specific geographical regions. One major challenge in designing effective interventions is to forecast age-specific overdose patterns to facilitate targeted prevention and preparedness efforts. To this end, we propose a forecasting approach that integrates observational data with an age-structured model of addiction and overdose mortality. Applying this method nationwide and in areas that are highly impacted by the overdose crisis, we provide robust drug-overdose mortality forecasts offering vital insights for effective interventions.

---

The United States is currently experiencing one of its worst drug crises, with alarming increases in fatal overdose rates. According to data from the Center for Disease Control (CDC), over 108,000 persons died from drug overdose in 2021, the highest number ever recorded in a single year and a 17% increase over the previous record high in 2020 [1]. Most recent overdose deaths involve synthetic opioids such as fentanyl, psychostimulants such as methamphetamines and, to a lesser degree, prescription opioids such as oxycodone, and heroin [2]. Many factors may have contributed to this surge, including the overall increased supply of synthetic, low-cost drugs [3, 4], the ease with which illegal substances may be purchased online [5–7], the uncontrolled mixing of drugs of different potency [8, 9], and societal changes leading to “deaths of despair” [10, 11]. Although these elements have fueled drug abuse for quite some time, most of them have been exacerbated by the COVID-19 pandemic [12, 13].

Both the CDC and the National Center for Health Statistics have been systematically collecting information on overdose mortality since 1999, and, according to slightly different classifications, since 1979. The relevant data is publicly accessible through the CDC Wide-ranging Online Data for Epidemiologic Research (WONDER) portal which is updated at the end of each calendar year with final data associated to the prior year, resulting in a one-year lag. Many groups have dissected these data by stratifying overdoses according to substance type, year, age, gender, race, and geography. These studies have revealed several spatio-temporal “overdose waves’ across the United States, the emergence of new trends, demographic and geographical shifts, and social disparities [13–16].

While providing up-to-date snapshots and following the course of past overdose deaths helps shed light on the evolution of the drug epidemic [17], forecasting future overdose patterns, even in the short term, would allow for targeted preventive interventions and ensure the preparedness of public health agencies [14, 18, 19]. Due to demographic, political and legislative heterogeneities across the United States, predictions on the national scale would be much less effective than those made at the more local level [20]. Analysis at a more “granular” scale retains specific drug-market, socioeconomic, cultural, and geo-historical conditions that distinctly affect the drug overdose trajectories. By not lumping these factors together, more realistic forecasts and tailored interventions [21, 22] can be developed. For instance, while drug overdoses may be decreasing at the regional level, certain counties, urban centers, or even zip codes within the same region may be experiencing surges among given sub-populations due to the introduction of new drugs to a circumscribed market.

As data collecting and manipulation capabilities have expanded, predicting overdose mortality (at any scale), while still in its infancy, has become a rapidly growing field. Given the many aspects of the drug addiction crisis, current studies rely on a variety of information including data on past overdoses, hospitalization, arrest, internet searches, painkiller prescription and drug-seizure rates. Quantitative tools used in these endeavors include statistical regression, geospatial analyses, mathematical modeling and machine learning [23–30].

In this paper we advance the state-of-the-art in drug overdose forecasting by combining a mechanistic model describing age-stratified drug-overdose fatalities with recorded mortalities using data assimilation techniques [31–33]. The latter were first developed within the geological and atmospheric sciences to merge high-dimensional dynamical systems with large datasets to produce weather and climate forecasts. After decades of continuous improvement to both algorithms and computing infrastructure, modern operational weather forecasting centers are able to process about 10^7^ observations per day [34]. In addition to its application in climate dynamics, data assimilation has been used to estimate parameters in systems biology [35], to provide risk-dependent individual contact interventions during outbreaks [36], to identify patients with antibiotic-resistant bacteria in hospital wards [37], and to quantify the proportion of undocumented COVID-19 cases [38]. One reason for the successful integration of mechanistic models with data assimilation methods across different fields is that the algorithms are computationally efficient and provide good forecasts even when training data are sparse [39]. Furthermore, since they are coupled to mechanistic models, data assimilation methods allow to estimate parameters that carry a physical or biological meaning and to follow their evolution over time. This interpretability, both of the parameters and of their dynamics, is very valuable for decision-making and formulating intervention policies. Finally, contrary to other techniques, data assimilation methods produce interval estimates and not just point estimates. This allows to quantify confidence intervals and accurately assess uncertainties and risks.

The mechanistic model that we use in this work is based the Kermack–McKendrick theory [40–44] and describes an age-structured population that suffers from substance use disorder (SUD). Our model includes population aging, addiction (*i*.*e*., the age-dependent onset of SUD in a certain subset of the overall population), and drug-induced mortality. Using data assimilation to combine our drug-overdose model with data from CDC WONDER, we develop a forecasting tool for age-stratified drug-overdose mortality in the United States. In the next section, we illustrate the basic principles of our method by generating nationwide drug-overdose mortality forecasts and by extracting the time evolution of epidemiological quantities such as mortality and addiction rates. We compare our predictions with overdose data for select past years and offer short-term projections for drug overdose mortality. Similarly, we generate drug-overdose forecasts for select counties or metropolitan areas that display a large number of overdose fatalities: Los Angeles County, CA, Cook County, IL and the five boroughs of New York City. Our forecasts show that age-structured population models combined with data assimilation methods can produce reliable predictions of drug-overdose deaths both at the national and county levels. Our approach and its results can inform early warning systems, help tailor interventions, and prioritize resources distribution to areas most impacted by the current drug epidemic.

## RESULTS

### Forecasting overdose fatalities in the United States

The Kermack–McKendrick model [40–43] is a standard tool in mathematical epidemiology and is used to describe the evolution of an age-structured population with age-dependent infection and recovery rates. Related structured population models have found utility in describing cell populations [45], demographics and birth control policies [46], the progression of infectious diseases [44] such as measles [47], tuberculosis [48], HIV [49], and COVID-19 [50] and more recently in the social sciences [51] and in studies of drug addiction [52–57]. In this work, we combine an age-stratified model of overdose fatalities with corresponding observational data from the CDC WONDER database, using an ensemble Kalman filter (EnKF) [58] as data assimilation method (see Online Methods for further information on the age-structured model, ensemble Kalman filter, and overdose data).

We model both the evolution of individuals with SUD and the number of fatal drug overdoses across different age classes in yearly increments. To estimate age-specific addiction rates (*i*.*e*., the influx of new SUD cases in a certain age class per unit time) and mortality rates, we use the age-structured population data and fatal overdose data tallied by the CDC WONDER database as inputs to our EnKF. Figures 1(a,b) show the evolution of the United States population and overdose fatalities from 1999 (light blue) to 2021 (dark blue). Within this timeframe, the population of the country between ages 0–85 rose from 275 to 326 million individuals; Figure 1(a) shows that the largest increases occurred between ages 20-40 years and that the age-structured population distribution is marked by two characteristic peaks: one arising between ages 20–30 years and the other between ages 40–60 years. The age-structured fatal overdose distribution in Fig. 1(b) reveals that between 1999 and 2014, the largest proportion of overdose deaths occurred between ages 40-50 years. During a second phase, spanning from 2015 to 2019, overdoses peaked within the 35-40 year age group. A sudden surge in overdose fatalities is observed beginning in 2020; the onset of this third phase is concurrent with the advent of COVID-19. These three phases do not define rigid classifications; rather, they provide reference points to facilitate data interpretation and guide our analysis.

**FIG. 1.**
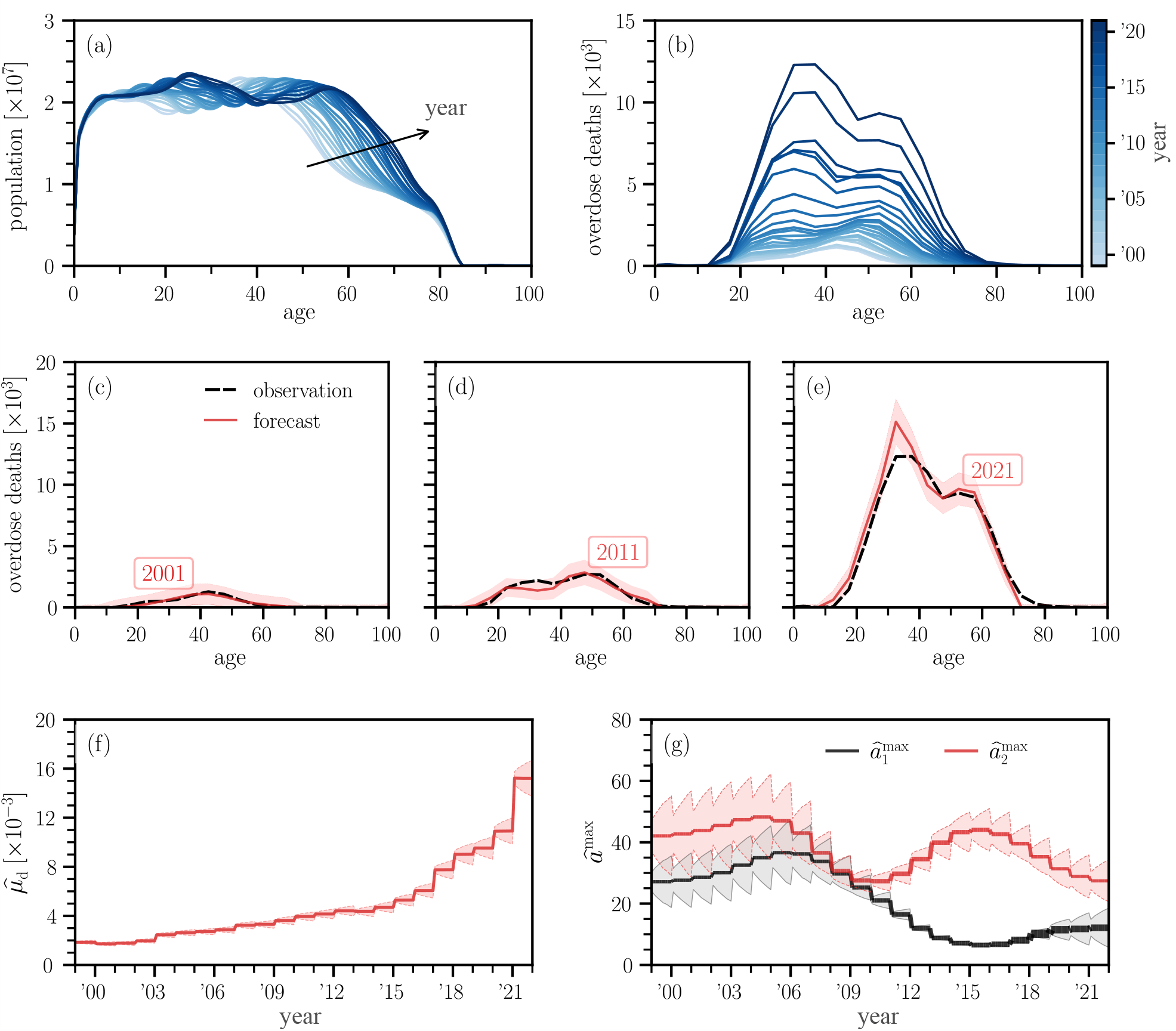
Forecasting nationwide overdose fatalities. (a,b) United States population and overdose deaths as a function of age (0–100 years) and time (1999–2021). (c–e) Forecasts of overdose deaths as a function of age. Solid red curves and red shaded areas indicate mean predictions and 3*σ* intervals, respectively. Observed fatalities are indicated by dashed black curves. Ages are binned in 5 year intervals and prediction values are displayed at the center of each bin interval. (f) Evolution of estimated drug-caused mortality rate 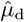 and corresponding 3*σ* intervals. (g) Evolution of the estimated ages for which the onset of SUD is largest and corresponding 3*σ* intervals. To account for potential population shifts, we utilize two age-stratified influxes peaked at 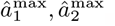. The thickness of the curves is proportional to their respective magnitudes 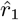 and 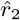. As can be visually inferred, the influx peaked at the younger age 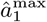 begins to carry more weight than its counterpart peaked at 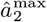 around 2015, indicating a shift towards a preponderance of young persons with SUD. Filter updates occur in the beginning of each year.

We use these qualitative observations to guide the development of our age-structured McKendrick model. First, to allow for possible population shifts or shifts in the onset of SUD, we include two age-stratified influxes of new SUD cases in the shape of gamma distributions peaked at ages 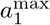 and 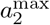 with amplitudes *r*_1_ and *r*_2_, respectively. Large values of *r*_1_ compared to *r*_2_ imply that the influx of new SUD cases occurs mostly through the distribution that is peaked at 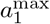 and vice-versa. Second, to take into account non-overdose deaths among drug users, the mortality rate we use for the population with SUD is given by an age-stratified baseline value derived from the Gompertz–Makeham–Siler mortality model [59–63] to which an excess drug-induced mortality *μ*_d_ is added.

As typical in data assimilation, at each forecasting step we use new overdose fatality data to update the system state and estimates of model quantities (such as the drug-induced mortality rate 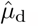, the ages at which the rate of the onset of SUD is maximal, 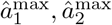, and the amplitudes of the SUD influx distributions 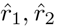) and use these values for subsequent forecasts. Since CDC WONDER data is available from 1999 until 2021, a forecast for drug-overdose fatalities in year *Y* is based on assimilated observational data between 1999 and *Y−*1. Figures 1(c–e) display age-stratified overdose forecasts (solid red curves) and corresponding observational data (dashed black curves) for the years 2001, 2011, and 2021. These are representative years selected from the three phases outlined above. In all panels red shaded regions indicate 3*σ* intervals and ages are binned in 5 year intervals. The shown nationwide forecasts exhibit a remarkable similarity to the actual observations; despite a significant rise in overdose fatalities between 2019 and 2020, the EnKF forecast of age-stratified overdose deaths remains in close agreement with the reported number of fatalities.

In addition to forecasting fatalities in each age group, we also used our EnKF to estimate the trajectories of 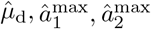 and 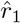 and 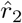 over the 1999–2021 interval. The EnKF was initialized with 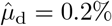 per year, consistent with 1999 data [64]; we also set the initial values 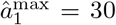 years, 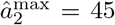 years and 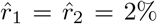 per year. Figure 1(f) shows that 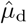 increased more than 7-fold in the past 20 years, rising from about 0.2% per year to 1.5% per year. This finding is largely independent of the initial value of 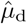; the final estimate, 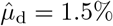 overdose deaths per year is larger than the baseline Gompertz–Makeham–Siler mortality rates for all ages under 60 years old.

The trajectory of the quantities 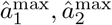 presented in Fig. 1(g) show that between 1999 and 2009 these values remain approximately stable within the 30–35 and 45–50 year range, respectively. In later years however, while 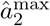 decreases only mildly, there is a strong descent of 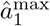 towards lower values, even below 20 years, indicating a substantial inflow of SUD cases at lower ages. Figure 1(g) also shows that 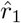 increases within the period of observation and that in 2015 it surpasses 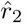, so that the influx of persons with SUD is dominated by the distribution peaked at the younger age 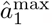. The shift of the onset of SUD towards younger ages that is observed starting in 2015 is consistent with the concurrent emergence of high mortality rates within the 25–35 age group as seen in Fig. 1(b).

Assessing the mortality rate of the population with SUD is challenging since a large number of subjects must be recruited and followed to evaluate the occurrence of a rare event such as death. Typical studies are conducted among SUD patients enrolled in treatment clinical trials or who have been hospitalized and monitored post-discharge. These studies reveal that drug users exhibit elevated mortality compared to the general population primarily due to fatal overdoses, but also due to viral infections, cardiovascular disease, and cancer [65, 66]. Overdose-specific mortality rates among SUD patients vary between 0.2% and 1% depending on gender, drug of choice, treatment type [66–69]; other meta-studies reveal that the mortality rate for persons who abuse opioids is about 0.7% [70]. Our estimates for 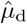 from Fig. 1(e) are in agreement with these values; in addition, our work enables the tracking of longitudinal changes in overdose mortality rates throughout the entire 1999–2021 period, uncovering a significant and alarming surge in mortality rates among individuals with SUD. Particularly noteworthy is the pronounced increase observed during the years 2020 and 2021.

Finally, in Fig. 2 we show nationwide forecasts for the years 2022, 2023, 2024 using our EnKF and the 2021 values of 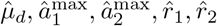. These are overdose fatality forecasts for which observational data are not available at the time of writing (Summer 2023); data for the year 2022 will only be made public by the CDC in early 2024. Our forecasts show that the number of overdose fatalities in the next few years will remain high and will continue to exceed those reported in 2019 for all age groups. Fatalities will remain largest among those younger than 30, with drug overdose counts matching the record values of the pandemic year 2021. However, we expect to see a decrease in overdose deaths in age groups older than 30.

**FIG. 2.**
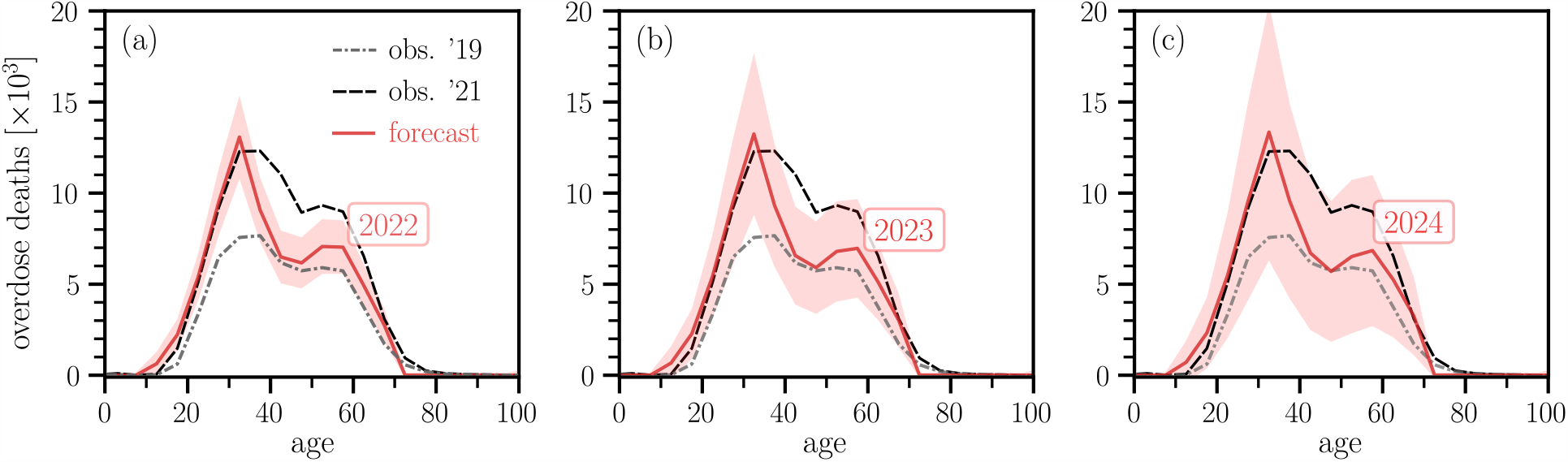
Forecasting nationwide overdose fatalities for 2022–2024. (a) Forecasts of overdose deaths in the United States as a function of age (0–100 years) for the year 2022. Data for 2022 is not available at the time of writing (Summer 2023) and will be made public by the CDC in early 2024. The forecast (solid red curve) is higher than the 2019 observations (dash-dotted gray curve) for all age groups but less than the 2021 data (dashed black curve) for ages greater than 30. Light red shaded regions show values within 3*σ*. The predicted number of overdose deaths will stay near the record highs recorded in 2021 among younger persons, until age 30. (b,c) Forecasts of overdose deaths in the United States as a function of age (0–100 years) for the years 2023 and 2024. Predicted trends are similar to 2022 projections and indicate that overdose fatalities for those under age 30 will remain high in the near future. As there is no available data for 2022 to 2024 the 3*σ* intervals increase due to the larger uncertainty as time progresses. Ages are binned in 5 year intervals and prediction values are displayed at the center of each bin interval.

To summarize, our findings reveal a staggering 7-fold increase in mortality rates among individuals with SUD, a generational shift towards drug use at younger ages, and alarming numbers of past and projected overdose deaths among individuals up to 30 years old. These results emphasize the necessity for more focused approaches in intervention and prevention strategies.

### County-level variation

Although the overall number of drug-overdose fatalities in the United States is rising, it varies significantly across counties. Figure 3(a) shows the distribution of county-stratified drug-overdose fatalities for select years between 2000–2020. Only counties with statistically significant fatalities of more than 10 individuals per year are shown. The number of counties that reached this significance threshold increased from 61 counties (out of 3, 147) in 1999 to 742 (out of 3, 142) in 2021, as reported in the CDC WONDER database. Between 1999 to 2021, numerous counties reported annual numbers of overdose fatalities below 100. However, during the same period, the number of counties experiencing between 100 and 1,000 annual overdose fatalities steadily increased. In 2020 (blue disks) a few counties even recorded close to 1, 000 overdose deaths. Crude rates, defined as the number of deaths per 100, 000 persons, also increased significantly over time, as seen in Fig. 3(b): in the year 2000 the mean crude rate among all counties for which data was available was 4.3 cases per 100, 000, in 2020 it was 31.5 cases per 100,000. This 7-fold increase is consistent with the similar rise in 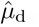 as inferred by our EnKF on the national level. The distributions in Fig. 3(b) also show that the crude rates exhibit a high degree of variability across counties.

**FIG. 3.**
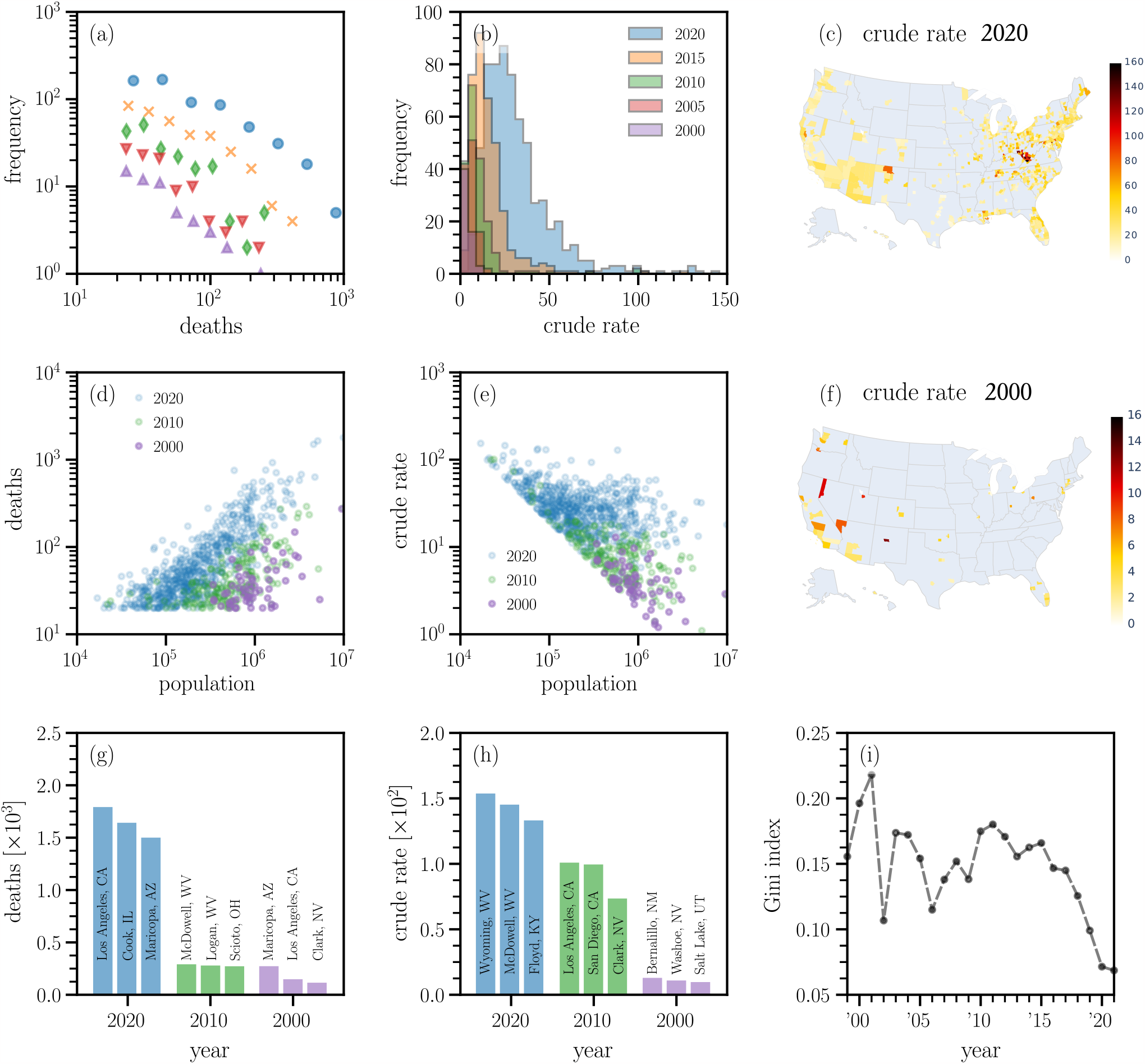
Drug-overdose fatalities in the United States at the county level. (a) Histogram of county-stratified drug-overdose fatalities for different years (blue disks: 2020, orange crosses: 2015, green diamonds: 2010, red inverted triangles: 2005, purple triangles: 2000). (b) Histogram of county-stratified crude rate data for different years. (d) County-stratified drug-overdose fatalities as a function of the corresponding county populations. (e) County-stratified crude rates as a function of the corresponding county populations. (c,f) Crude rates across different counties in 2020 and 2000. The scales of the two color bars differ by a factor of 10. In the gray regions, either no data or a statistically not significant number of cases were reported. In all panels, we did not include data for which at least one database entry (*e*.*g*., fatalities and crude rate) was marked unreliable. The minimum number of deaths in the remaining data is 20. Hence, the crude rate data in (e) lies above of the minimum crude rate given by 2*×* 10^6^*/*Population. (g) The three counties with the largest overdose death tolls in 2020, 2010, and 2000. (h) The three counties with the largest overdose crude rates in 2020, 2010, and 2000. (i) The Gini index across different years. A Gini index of 0 means that the crude rate is the same across all counties. If all overdose fatalities were concentrated in one out of *N*_c_ counties, the Gini index would be 1*−*1*/N*_c_. The number of counties with statistically significant fatality counts (greater than 10 deaths in a given year) and crude rates are *N*_c_ = 61 in 1999 and *N*_c_ = 742 in 2021.

Figure 3(d) shows that in 2000, only counties with populations larger than 100, 000 residents experienced statistically significant numbers of drug-overdose fatalities. In the years since, crude rates substantially increased for these counties, especially between 2010 and 2020, while smaller counties with population sizes of about 10, 000 also started reporting significant numbers of fatal overdoses, as shown in Fig. 3(e). This indicates that the drug overdose epidemic has permeated all jurisdictions, regardless of population.

The heat maps in Figs. 3(c,f) confirm the increase in the number of counties affected by the drug epidemic between 2000 and 2020. Notice that the scales of the two color bars differ by a factor of 10. Figure 3(c) shows that in 2000, the most affected areas were population centers in the Western United States: that year, the largest overdose fatality counts occurred in Maricopa County, AZ; Los Angeles County, CA; and Clark County, NV [Fig. 3(g)] and the largest crude rates were reported in Bernalillo County, NM; Washoe County, NV and Salt Lake County, UT [Fig. 3(h)]. In 2020, many regions in the Central and Eastern United States also became heavily impacted by the drug epidemic, including many smaller population counties [Fig. 3(f)]. The largest 2020 fatality county were reported in Los Angeles County, CA; Cook County, IL; and Maricopa County, AZ [Fig. 3(g)]. The largest 2020 crude rates were registered in Wyoming County, WV; McDowell County, WV; and Floyd County, KY [Fig. 3(h)].

In 2000, Los Angeles County, the most populous in the United States, accounted for approximately 12% of the total population of the 59 counties with statistically significant overdose fatalities. Its proportion of overdose fatalities was about 10%. However, due to more counties reporting large numbers of overdose deaths, by 2020, Los Angeles County’s population represented only 4% of the total population of the 640 counties with statistically significant overdose deaths. That same year, Los Angeles County contributed to approximately 3% of the registered overdose deaths at the county level. Furthermore, in 2020, despite the 10 most populous counties in the United States being home to roughly 16% of the population (out of the 258 million associated with the 640 counties with statistically significant overdose fatalities), they recorded less than 13% of the number of overdose fatalities among these 640 counties. Conversely, the 50 least populous counties, with less than 1% of the total population among the 640 affected counties, accounted for more than 2% of the number of overdose deaths. These statistics highlight the shifting patterns of overdose fatalities, with a notable rise of overdose deaths in less populated counties. To better understand how the drug deaths are shared across counties, we consider the Gini coefficient [71, 72], a measure of inequality among a set of *N*_c_ values of a distribution; in our case *N*_c_ is the number of counties that have reported statistically significant numbers of overdose deaths. We compute the Gini index by plotting the proportion of the total number of overdose fatalities accumulated across counties against the cumulative population fraction across counties. The lower bound for the Gini index is 0 (perfect equality, indicating that overdose deaths and county populations are proportional), and the upper bound is 1*−*1*/N*_c_ (perfect inequality, indicating that all overdose deaths occurred within a single county). We find that the Gini coefficient dropped from a value of about 0.2 in the year 2000 (*N*_c_ = 59), to about 0.07 in the year 2021 (*N*_c_ = 640), which is consistent with increases in the number of counties affected by the drug-overdose epidemic. In the next section, we employ the modeling and forecasting techniques established in the previous section to examine the progression of age-specific overdose fatality counts in three specific regions: Los Angeles County, CA; Cook County, IL; and the combined area of New York City’s five boroughs (The Bronx, Brooklyn, Manhattan, Queens, and Staten Island).

### Forecasting overdose fatalities in three counties

The dynamics of overdose fatalities in Los Angeles County, Cook County and five boroughs of New York City have unfolded in substantially different ways over the past two decades. In 1999, Cook County reported a total of 14 overdose fatalities, New York City recorded 52, and Los Angeles County 384. By 2021, these numbers had risen to 1,699 overdose deaths for Cook County, 2,091 for Los Angeles County, and 2,124 for New York City. Cook County experienced the most striking rise in overdose fatalities between 1999 to 2021: an unprecedented 120-fold increase. In comparison, New York City experienced a 40-fold increase and Los Angeles County a 5-fold increase. These numbers are even more striking given Cook County’s smaller population (5.1 million in 2021), compared to the population of Los Angeles County (9.7 million in 2021) and New York City (8.3 million in 2021).

As in our nationwide forecasts, we use an EnKF in conjunction with an age-structured overdose mortality model that accounts for the underlying age variation in the county populations. Because of the relatively small overdose fatality counts in Cook County and New York City in the early 2000s, we do not report forecasts for 2001 as done nationwide and for Los Angeles County, but we use years from 2013 onwards for which enough data is available across all age groups. Specifically, in Cook County, the total number of reported age-stratified drug-overdose fatalities remained below 100 for most years prior to 2013. This points to a delayed emergence of the overdose death crisis in this county which became extraordinarily acute in just a few years. A finer analysis reveals that the largest increases in drug overdose mortality in Cook County are due to heroin (a 10-fold rise between 2012 and 2013) and to fentanyl (a 5-fold rise between 2015 and 2016).

Results are shown in Fig. 4. The first three historical forecasts for each region are compared to the corresponding observational data (dashed black curves). Since the observational data for 2023 is not available at the time of writing (Summer 2023), we display the 2019 data (dash-dotted gray curve) and the 2021 data (dashed black curve) in the 2023 forecast. For Los Angeles County, we initialized the EnKF with 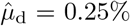 per year and 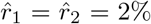 per year. The simulations for Cook County and New York City start in 2013, and we initially set 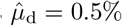 per year and 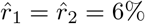 per year. The estimated drug-caused mortality rate for Cook County and New York City in 2021 are, respectively, 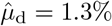 per year and 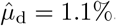, substantially larger than the 2021 estimates for Los Angeles County (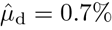 per year). This is also consistent with the much larger increase in overdose fatalities in Cook County and New York City compared to Los Angeles County.

**FIG. 4.**
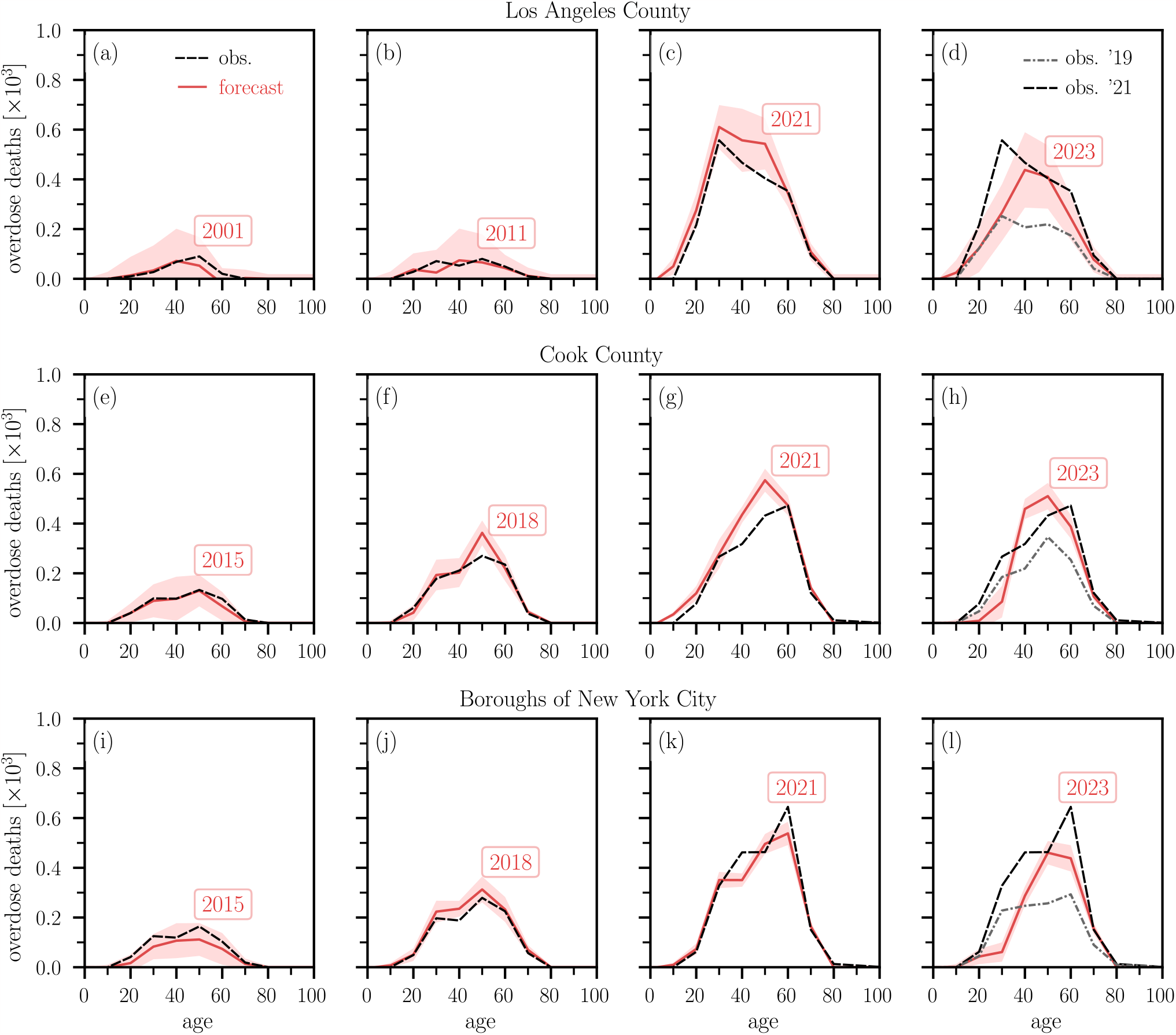
Forecasting overdose fatalities in select US jurisdictions. (a–d) Forecasts of overdose deaths in Los Angeles County as a function of age (0–100 years) for 2001, 2011, 2021, and 2023. Solid red curves and shaded regions indicate mean predictions and 3*σ* intervals, respectively. Observed fatalities are indicated by dashed black curves. Because observational data for 2023 is not available at the time of writing, we show 2019 data (dash-dotted gray curve) and 2021 data (dashed black curve) in panels (d,h,l). (e–h) Forecasts for Cook County using the same graphic representations as in panels (a–d) and for the years 2015, 2018, 2021, 2023. (i–l) Forecasts for the five boroughs that comprise New York City (The Bronx, Brooklyn, Manhattan, Queens, and Staten Island) using the same graphic representations as in panels (a–d) and for the years 2007, 2014, 2021, 2023. Although the numerical escalation in drug-overdose deaths in Cook County (2015–2021) and New York City (2007–2021) is similar to what is observed for Los Angeles County, the timelines are much accelerated. In 2021 the population of Los Angeles County was 9.7 million, in Cook County 5.1 million and in New York City 8.3 million, indicating a more acute crisis in Cook County. Ages are binned in 10 year intervals and prediction values are displayed at the center of each bin interval.

## DISCUSSION

Escalating drug-induced deaths have been a major public-health challenge in the United States for more than a century. The over-prescription of morphine and opium led to an epidemic that affected almost 5 in 1,000 Americans in the 1890s [73]. This widespread crisis spurred a number of acts and regulations in the early 20^th^ century that succeeded in lowering opiate use and mortality rates [74, 75]. The current epidemic involves a significantly higher prevalence of SUD and has unfolded via distinct spatio-temporal mortality waves driven by different drug types and localized sub-epidemics. Being able to forecast the complex evolution of fatal drug overdoses at the national, regional or county levels, would represent major advancements in helping curb drug abuse.

In this work, we developed a forecasting method that combines an age-structured model of addiction and overdose mortality with observational data derived from the CDC WONDER database through a data assimilation approach. By applying our method to nationwide data as well as to three representative areas (Los Angeles County, Cook County, and the five boroughs of New York City) we showed its ability to provide near-term forecasts, to extract epidemiological parameters, and to capture the heterogeneity in overdose mortality across different counties. Since the demographics and geography of drug abuse are in constant flux, we believe our data assimilation approach holds promise for informing targeted prevention and preparedness interventions aimed at curbing drug overdose deaths.

The nationwide drug-induced mortality rate has risen more than 7-fold in the past two decades, surpassing 1.5% among persons suffering from SUD. This rate exceeds the baseline Gompertz–Makeham–Siler mortality rates for all groups under 60 years old. Our county level analysis reveals significant variations in overdose fatality trends. For example, although at the onset of 1999–2021 period, Los Angeles County had a higher drug-induced mortality rate than Cook County and New York City, in 2021 it had the lowest, at 0.7%, compared to Cook County at 1.3% and New York City at 1.1%. Furthermore, the annual number of overdose deaths in Cook County and New York City grew much faster than in Los Angeles County in recent years. This points to a delayed, yet severe growth of drug overdoses in New York City and especially in Cook County. For past years, our predictions are in good agreement with tallied data. For the year 2023 we predict slight decreases of drug overdoses compared to the pandemic year 2021, both nationwide and in the counties we surveyed. Specifically, we expect drug overdose deaths to decrease nationwide by 17% compared to the values recorded in 2021, by 24% in Los Angeles County, by 32% in New York City but only by 8% in Cook County.

We also find that the overdose epidemic has spread to more counties over time. In the year 1999, 61 counties out of 3, 147 had statistically significant overdose fatalities, whereas this number rose to 742 out of 3, 142 in 2021. Not only has the number of affected counties grown, but their relative contributions to the overall overdose fatality count have become more evenly distributed over the years. This finding implies that managing the overdose epidemic cannot be simply accomplished by targeting a few specific counties, rather each jurisdiction must develop specific plans tailored to their unique socio-demographic and economic profiles.

Several limitations of this study are noteworthy. Our findings are based on four drug categories with the highest crude rates available in the CDC WONDER database: fentanyl (T40.4), prescription opioids (T40.2), heroin (T40.1), and methamphetamines (T43.6). We did not include other categories such as T40.3 (methadone) or T40.5 (cocaine) in our analysis due to their lower mortality rates. The dynamics of fatalities associated with these drug categories may differ from the fatality trends observed in our analysis. Furthermore, in certain jurisdictions, fatality data is unavailable as the CDC WONDER portal suppresses entries where the number of deaths is less than 10. Additionally, some overdose cases may involve multiple drugs. In such instances, deaths are counted in all relevant categories, resulting in multiple counts. Finally, comparisons of opioid-related overdose death rates at the national, state, and county levels may be influenced by significant variations in the reporting of specific drugs involved in overdose deaths. Changes in drug reporting specificity over time and across different states and counties can lead to potentially misleading conclusions regarding actual drug-specific death rates [76].

There are several potential avenues for future work. While we perform yearly parameter updates and forecasts using final data from CDC WONDER, it is also possible to utilize provisional fatality data, which are released at earlier dates and on a monthly basis, to generate more timely but similarly provisional forecasts; retrospective updates can then be performed once final data becomes available. Although we only analyzed three large urban areas, our method can be applied to other jurisdictions. In less populous areas the number of fatalities may not be sufficiently large for a meaningful age-stratified analysis; in these cases pooling data from several neighboring jurisdictions with similar socio-economic characteristics, using larger age-binning or considering biannual forecasts may yield more meaningful results. Alternatively, for small-number cases, a stochastic version of the Kermack–McKendrick model may be used as to evolve the state variables probabilistically [77]. Our methods can also be used to forecast regional drug-overdose mortality by gender, race or drug-type; the resulting projections for specific groups of users or drugs of abuse may help guide more effective intervention efforts. Large deviations of observed data from our projections would signal fundamental changes to the illicit drug landscape in the form of effective prevention and treatment programs, or in the consumption of more addictive or lethal substances. Finally, one may also study the numerical stability and forecasting accuracy alternative ensemble-based Kalman filters, such as ensemble adjustment Kalman filters [78], or incorporate backward passes and smoothing techniques into our method to potentially enhance earlier parameter estimates [79].

## Data Availability

All data produced are available online at the repositories mentioned in the manuscript.

## ACKNOWLEDGEMENTS

TC and MRD acknowledge financial support from the ARO through grant W911NF-18-1-0345; LB and MRD acknowledge financial support from the ARO through grant W911NF-23-1-0129.

## DATA AND CODE AVAILABILITY

Our source codes are publicly available at https://gitlab.com/ComputationalScience/overdose-da. The CDC WONDER portal can be accessed at https://wonder.cdc.gov/.

## METHODS

### Age-structured overdose model

The mathematical model we use to describe the age-stratified evolution of SUD cases is given by

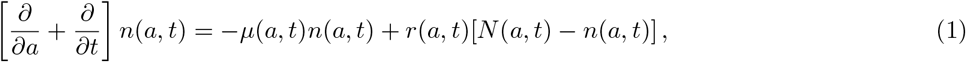

where *n*(*a, t*)d*a* is the population affected by SUD with age between *a* and *a* + d*a* at time *t*. The associated mortality is *μ*(*a, t*) and *r*(*a, t*) is the influx rate of new SUD cases from *N* (*a, t*)−*n*(*a, t*), the pool of individuals without SUD. Finally, *N* (*a, t*) is the general population with age between *a* and *a* + d*a* at time *t*. We set the initial age and time *a*_0_ = *t*_0_ = 0. The initial distribution of SUD cases is given by *n*(*a, t* = 0) = *ρ*(*a*). We also set *n*(*a* = 0, *t*) = 0 such that no population of age *a* = 0 exists at any time. We solve Eq. (1) using the method of characteristics and distinguish the two cases *a≥t* and *a < t*. For *a≥t*, the characteristic begins at *t* = 0 and *n*(*a, t*) will remain constant along *a* = *t*, yielding

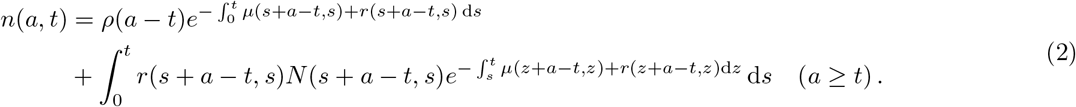

For *a < t*, the characteristic will begin at *a* = 0 and *n*(*a, t*) will remain constant along *t* = *a* so that

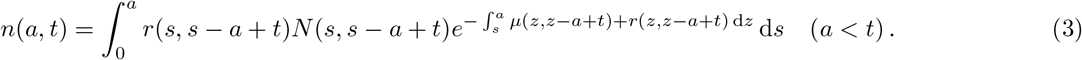

We write the mortality rate *μ*(*a, t*) as the sum of a baseline mortality rate, *μ*_0_(*a, t*), and a drug-caused excess mortality rate, *μ*_d_(*a, t*), so that *μ*(*a, t*) = *μ*_0_(*a, t*) + *μ*_d_(*a, t*). The baseline mortality *μ*_0_(*a, t*) = *μ*_0_(*a*) is derived from the Gompertz–Makeham–Siler mortality model for human death [59–63] and is assumed to be time-independent, implying that age-stratified mortality has not changed appreciably over the past twenty years. The quantity *μ*_d_(*a, t*) = *μ*_d_ is instead assumed to be age-independent and inferred from data, so that we effectively neglect any time dependence over a data assimilation cycle of one year. Within a data assimilation window of one year, we thus set

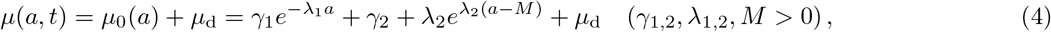

where *γ*_1_ = 0.00258*/*year, *γ*_2_ = 0.00037*/*year, *λ*_1_ = 5.09657*/*year, *λ*_2_ = 0.09040*/*year, and *M* = 83.22956 year in accordance with Ref. [63]. Since the majority of drug overdose fatalities occur among males [13, 80] as seen in Fig. 6 we explicitly used parameters pertaining to males in the United States. Parameters vary slightly from year to year, we selected the ones for 2010. It is worth noting that we conducted a sensitivity analysis through additional simulations using variations on the above choices for *γ*_1_, *γ*_2_, *λ*_1_, *λ*_2_, *M* including values specific to females in the United States. We find that the choice of these parameters does not substantially affect our results.

**FIG. 5.**
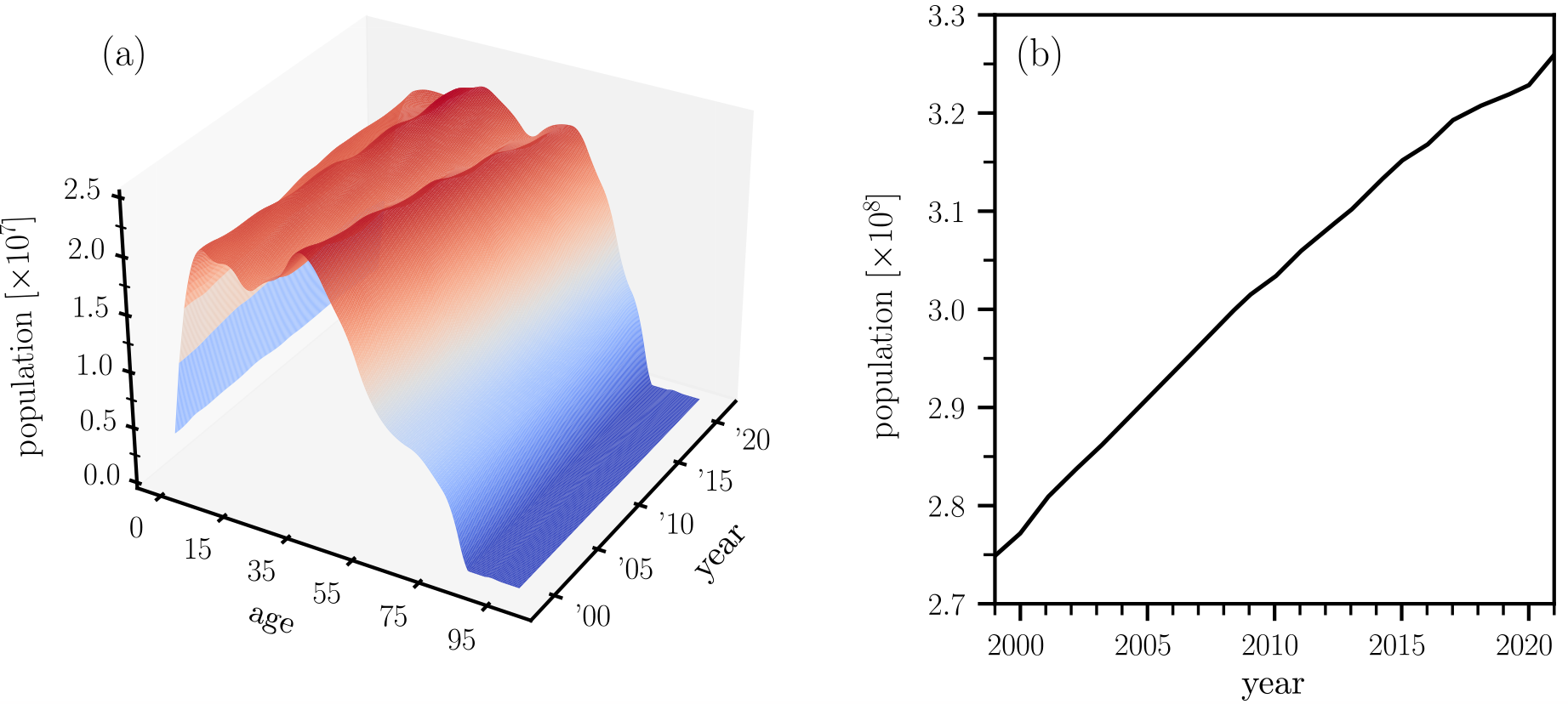
Age-structured United States population data. (a) Interpolated age-structured population data. The interpolation is based on bivariate splines of degree 2. (b) The overall United States population increases almost linearly between 1999 and 2021.

**FIG. 6.**
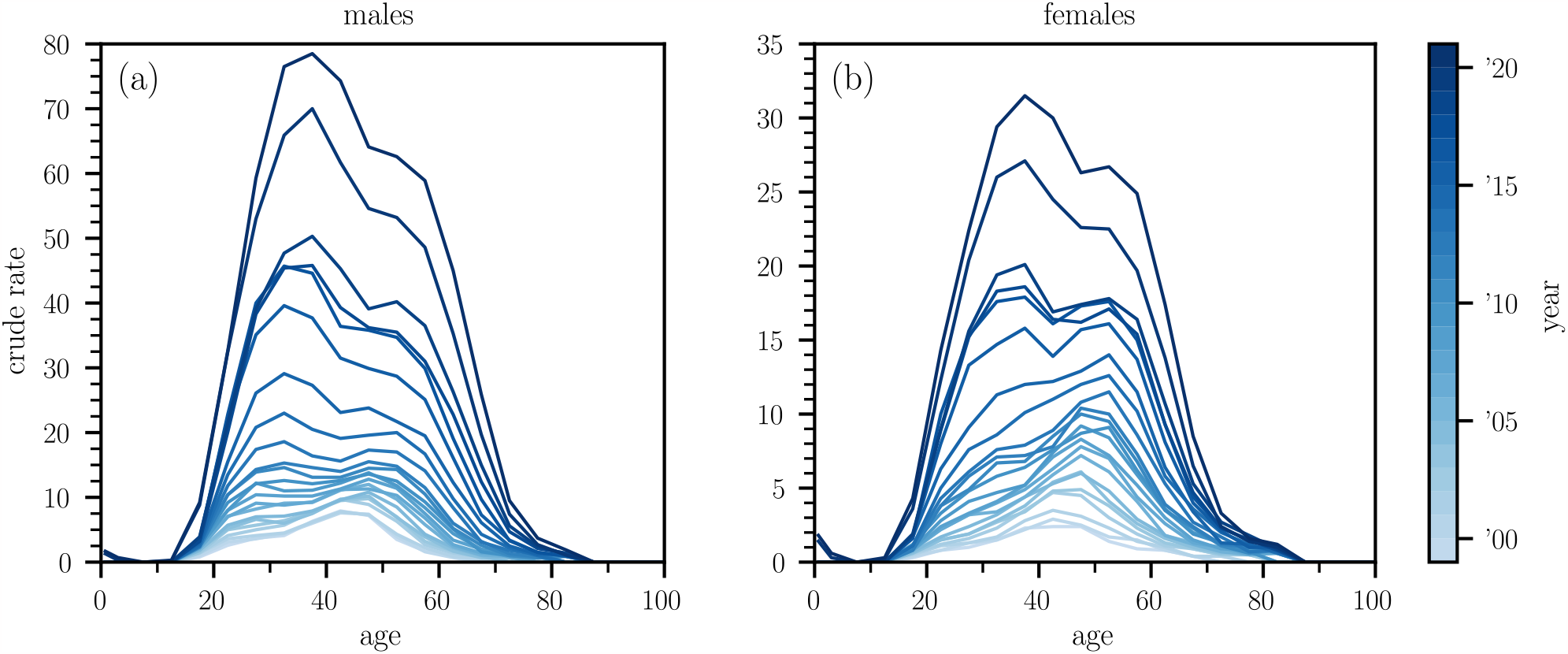
Age-structured overdose crude rates in the United States. Age-structured overdose crude rates in the United States, distinguished by gender: (a) males and (b) females.

We allow the parameter *μ*_d_ to change from one year to the next. This choice is dictated by the CDC WONDER database providing yearly lists of overdose deaths, although monthly updates could also be implemented. To describe an age-dependent influx into the pool of SUD users, we set *r*(*a, t*)*≡r*(*a*) = [*r*_1_*f* (*a*; *α*_1_, *β*_1_) + *r*_2_*f* (*a*; *α*_2_, *β*_2_)] */*2, where *f* (*a*; *α, β*) = *β*^*α*^*/*Γ(*α*) *a*^*α−*1^*e*^*−βa*^ is the gamma distribution with shape and rate parameters *α* and *β*. The maximum of a gamma function is given at the age *a*^max^ = (*α−*1)*/β*. The evolution of *n*(*a, t*) in data assimilation cycles requires us to evaluate the derivative of Eq. (2) w.r.t. *t*. Given the two variable function *g*(*x, y*), we use the notation *g*^*′*^(*x, y*) and *g*_*′*_(*x, y*) to indicate a partial derivative in the first and second argument of *g*, respectively. For *a t*, the rate of change of *n*(*a, t*) can thus be written as

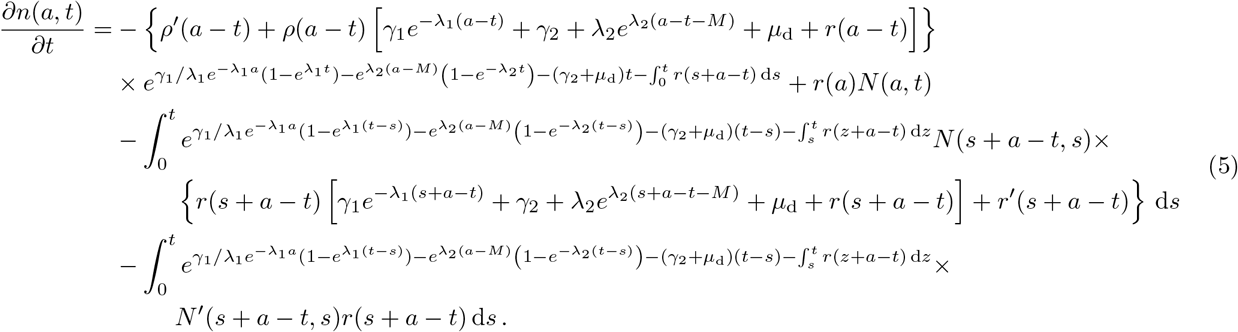

For *a < t* we obtain

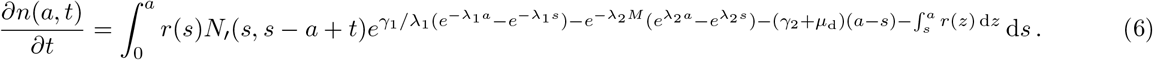

The integrals 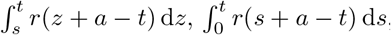, and 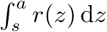 can be evaluated using the identity

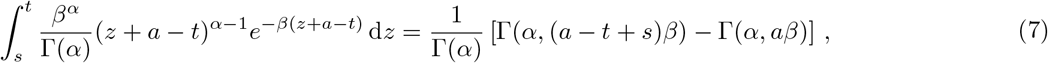

where 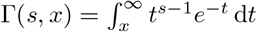 denotes the upper incomplete gamma function. We evaluate the remaining integrals 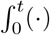 d*s* numerically. Finally, the initial condition used to solve Eq. (1) and to obtain the simulation results in Figs. 1, 2, and 4 is

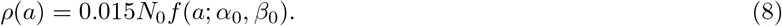

To obtain the curves shown in Figs. 1 and 2 we set *N*_0_ = 274, 886, 150, the population of the United States between ages 0–85 in 1999. We also select *f* (*a*; *α*_0_, *β*_0_) to be a gamma distribution with shape and rate parameters *α*_0_ and *β*_0_, chosen as *α*_0_ = 12 and *β*_0_ = 1*/*(3 year) such that the maximum of the distribution is at *a*_max_ = 33 years. The prefactor of 0.015 is chosen such that initially 1.5% of the population are suffering from an SUD consistent with corresponding survey data [64, 81]. To obtain the curves in the county-level analysis, we initially set *N*_0_ to match the respective population sizes between ages 0–85 for the years 1999 (Los Angeles County) and 2013 (Cook County and New York City). Specifically, *N*_0_ was set to 9, 437, 290 for Los Angeles County, 8, 405, 837 for New York City, and 5, 240, 700 for Cook County. We also set *α* = 17 (Los Angeles County) and *α* = 12 (Cook County and New York City) and used the same value of *β* as in the national analysis.

### Interpolating population data

We infer the age-structured population function *N* (*a, t*) from nationwide population data that is available from the CDC WONDER database. In Fig. 5(a) we show an interpolated and differentiable population function *N* (*a, t*). In all simulations, we use interpolations that are based on bivariate splines of degree 2. In Fig. 5(b), we show the almost linear increase of the population of the United States between ages 0-85 from 1999 to 2021.

### Ensemble Kalman filter

To combine the age-structured drug overdose model (1) with corresponding observational data, we use an ensemble Kalman filter (EnKF) [58] as implemented in [33, 83]. Figure 7 shows a schematic of the main EnKF steps.

**FIG. 7.**
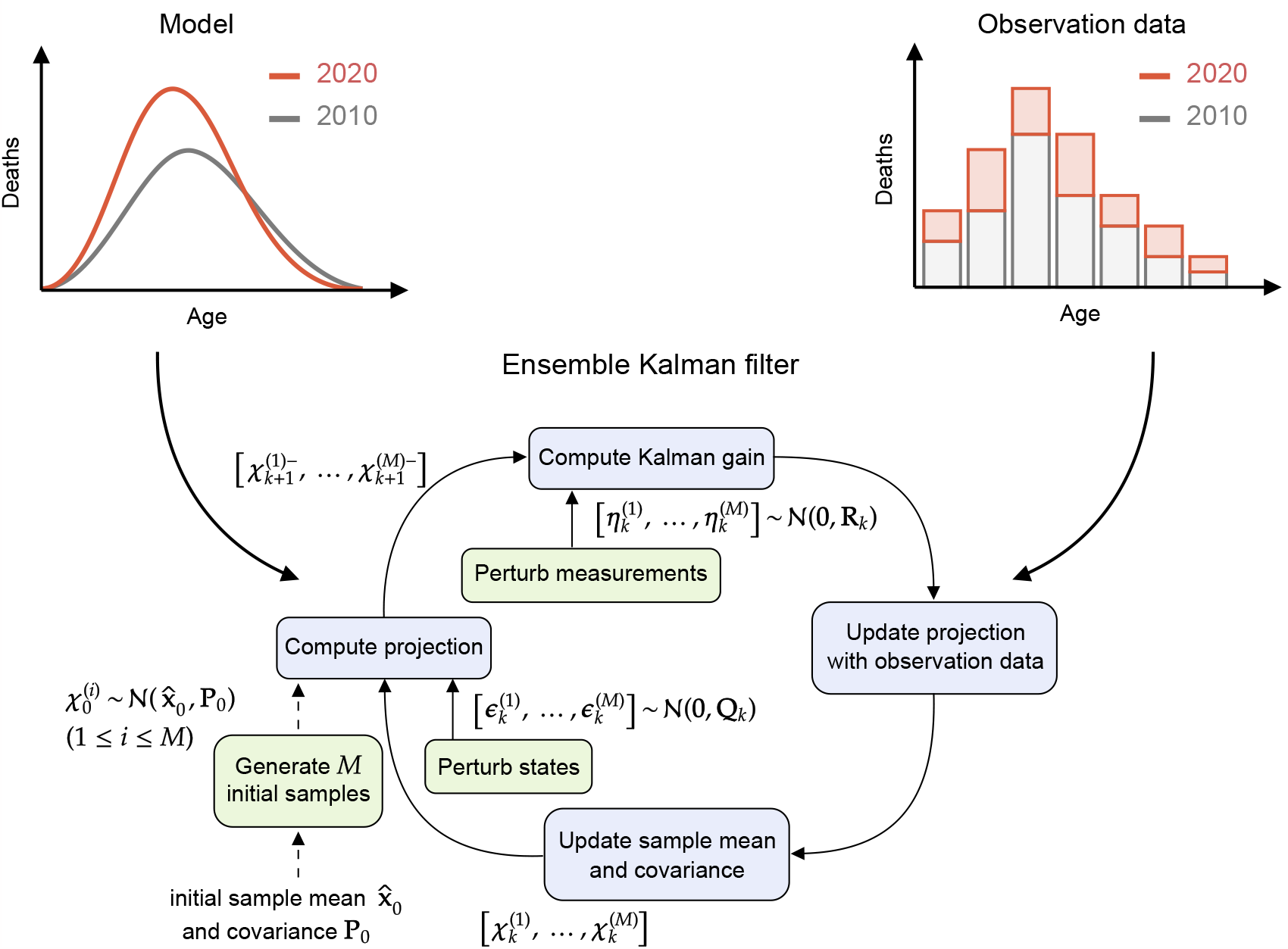
Ensemble Kalman filter schematic. We use an EnKF to combine a mechanistic model of drug-overdose fatalities with corresponding observational data. Blue boxes show the main steps (*i*.*e*., projection and update) in an EnKF cycle. Green boxes represent the initial sample generation process and perturbations that are added during the projection and update steps. The schematic is adapted from [82].

In accordance with [31, 84], the evolution of any system state **x**(*t*) (*e*.*g*., number of SUD cases, mortality and addiction rates) and observed state **z**(*t*) (*e*.*g*., number of overdose fatalities) are described by the stochastic differential equations

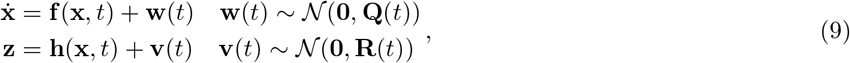

where **Q**(*t*) and **R**(*t*) denote the covariance matrices associated with the Gaussian process noise 𝒩 (**0, Q**(*t*)) and Gaussian measurement noise 𝒩 (**0, R**(*t*)) at time *t*, respectively. We assume the quantities **Q**(*t*) and **R**(*t*) to be given. The function **f** (·) describes the dynamics of the system state **x**(*t*), while **h**() maps **x**(*t*) to a measurable quantity. Both functions can be non-linear.

For the specific case of our age-structured model defined in Eq. (1), element *x*_*j*_(*t*) of the state vector **x**(*t*) corresponds to *n*(*a*_*j*_, *t*)*≡n*(*a*_0_ + (*j−*1)Δ*a, t*) (*j∈ {*1, …, *N*_*a*_*}*), the density of individuals whose age lies within the [*a*_0_ + (*j−*1)Δ*a, a*_0_ + *j*Δ*a*) interval at time *t*. Here, *N*_*a*_ and Δ*a* denote the number of discretizations of the age interval and the corresponding age discretization step, respectively. Thus, we write

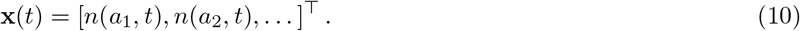

For the numerical solution of Eq. (9), we also discretize the simulation time interval [0, *T*] into *N*_*t*_ equidistant intervals of duration Δ*t* = *T/N*_*t*_. In all of our simulations, we fixed Δ*t* at 0.1. To combine the mechanistic model in Eq. (1) with empirical data on overdose fatalities, we augment the system state (10) by

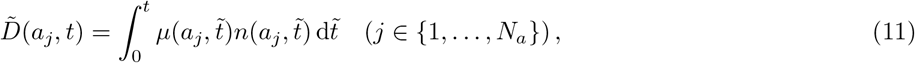

where 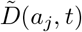 is the cumulative number of overdose deaths in the age interval [*a*_*j−*1_, *a*_*j*_) up to time *t*. To avoid dealing with large differences between predicted and observed fatalities in our numerical calculations, we normalize both quantities by dividing them by 1,000, thereby measuring overdose deaths per 1,000 individuals in the system state. Since we wish to estimate model parameters such as *μ*_d_ and *r*_1_, *r*_2_, *α*_1_, *β*_1_, *α*_2_, *β*_2_, we also augment the system state (10)by the log-transforms 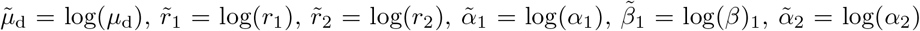 and 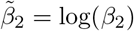. Therefore, the final augmented system state is

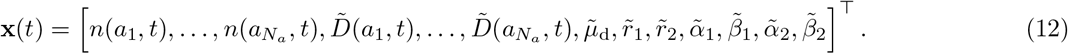

Prior to each prediction step, we apply an exponential transform to render the parameters *μ*_d_ and *r*_1_, *r*_2_, *α*_1_, *β*_1_, *α*_2_, *β*_2_ positive and avoid sign changes. To accurately solve the evolution of *n*(*a, t*) numerically, we must use a sufficiently large number of age windows *N*_*a*_ in our simulations. However, since the age windows in our simulations are more granular than those available from overdose fatality data, we apply a coarse-graining procedure. The nationwide CDC WONDER data we utilized has 22 age groups with 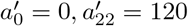 and 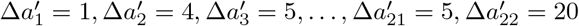 years. To differentiate between the age discretization in the observational data and the age discretization in the underlying model, we employ a superscript notation. In the county-level forecasts, we employed 10-year age groups, effectively reducing the number of age groups from 22 to 11. The following discussion of the EnKF parameterization and implementation will be based on 22 age groups, but the same considerations also apply to the county-level analysis with fewer age groups.

To reduce granularity and combine the modeled quantities 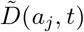 with corresponding observational data, we numerically integrate 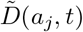 over the age windows 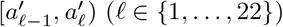 to obtain the cumulative number of deaths 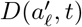 in this age interval at time *t*. Here, 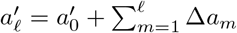 for *f ≥* 1. Based on the described mapping of 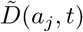 to 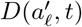, the measurement function becomes

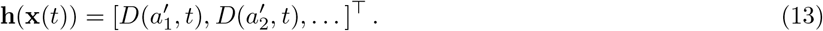

In our simulations, we set the initial values 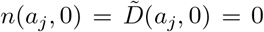. In the nationwide analysis, we initially set *μ*_d_ = 2*×*10^*−*3^*/*year, *r*_1_ = *r*_2_ = 2*×*10^*−*2^*/*year, *α*_1_ = 10, *β*_1_ = 1*/*(3 year), *α*_2_ = 15, and *β*_2_ = 1*/*(3 year). We have chosen the initial values of *r*_1_, *r*_2_ in accordance with corresponding empirical data on the number of substance initiates [64, 81] The initial maximum values of the gamma distributions *f* (*a*; *α*_1_, *β*_1_), *f* (*a*; *α*_2_, *β*_2_) are attained at ages 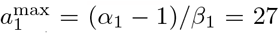 years and 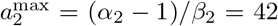 years, respectively. All initial covariances are set to 10^*−*4^, except for the diagonal elements associated with the log-transforms of *μ*_d_, *r*_1_, *r*_2_ and *α*_1_, *β*_1_, *α*_2_, *β*_2_, which are set to 1, respectively. Process and observation noise covariances are assumed to be time-independent and given by 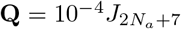 and **R** = diag(2*×*10^*−*3^, …, 2*×*10^*−*3^), respectively. Here, *J*_*n*_ denotes the *n×n* matrix of ones. Our simulations run from the beginning of 1999 until the end of 2024. The age discretization is Δ*a* = 1.2 years with *a*_0_ = 0 and *aN*_*a*_ = 120 years. Population data is available for ages between 0 and 85. However, since overdose fatalities in groups below 10 years and above 70 years are statistically insignificant, we truncated the system state accordingly. To align model parameters with initial observational data, we performed two full data assimilation cycles for the first year (1999) before starting the main forecasting algorithm that produces forecasts for all years from 1999 to 2024. The number of EnKF ensemble members is *M* = 10^3^.

We used different initial values for the county-level forecasts. For Los Angeles County, the initial values were set as follows: *μ*_d_ = 2.5*×*10^*−*3^*/*year, *r*_1_ = *r*_2_ = 2*×*10^*−*2^*/*year, and *α*_1_ = *α*_2_ = 17. For Cook County, the values were set as *μ*_d_ = 5*×*10^*−*3^*/*year, *r*_1_ = *r*_2_ = 6*×*10^*−*2^*/*year, *α*_1_ = 8, and *α*_2_ = 15. Lastly, for New York City, the values were set as *μ*_d_ = 5*×*10^*−*3^*/*year, *r*_1_ = *r*_2_ = 6*×*10^*−*2^*/*year, *α*_1_ = 8, and *α*_2_ = 17. The initial values of *β*_1_ and *β*_2_ are as in the nationwide analysis. To account for the smaller number of overdose fatalities at the county level, we adjust the process and observational noise matrix elements to have values of the order 10^*−*8^*−*10^*−*7^. The number of EnKF ensemble members is *M* = 10^3^ (Los Angeles County) and *M* = 10^2^ (Cook County and New York City).

At every time point *t*, we use the EnKF to determine the state posterior distribution given all prior observations. Before starting the data assimilation procedure, we generate an initial ensemble 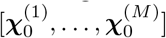 that consists of *M* ensemble members 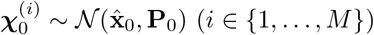. The quantities 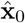 and **P**_0_ denote the given initial state and covariance estimates, respectively.

To perform forecast and update iterations using a Kalman filter, one uses state estimates 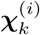 at time *t*_*k*_ to calculate predicted state estimates 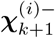 at time *t*_*k*+1_. These predicted state estimates are then combined with observational data to obtain an updated state 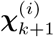 . We use the superscript “*−*” in 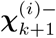^*−*^ to distinguish between predicted (*i*.*e*., prior) state estimates and updated (*i*.*e*., posterior) state estimates. In the remainder of this section, we describe the two main EnKF steps: (i) forecasting the evolution of the system state and (ii) updating the predicted state estimates using observational data. We use the shorthand notation *y*_*k*_ *≡ y*(*t*_*k*_) to refer to a quantity *y* at time *t*_*k*_ = *k*Δ*t* (*k ∈ {*0, …, *N*_*t*_*}*).

1. **Forecast Step**: For each ensemble member, the predicted state estimate 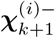 at time *t*_*k*+1_ is given by

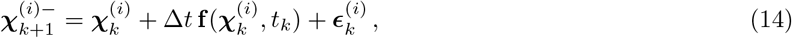

Where 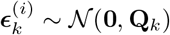 models Gaussian process noise. Using the predicted state estimates 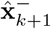, we compute the corresponding ensemble mean, 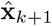, and covariance matrix, 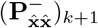, according to

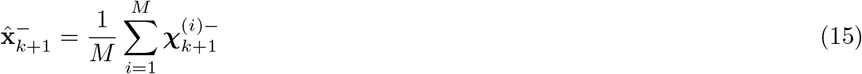

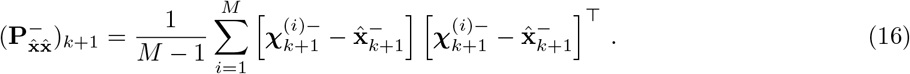

Although the covariance matrix 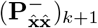 is not required in the EnKF iteration, it is useful to estimate confidence intervals of 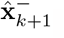.
2. **Update Step:** We first compute the ensemble mean of the predicted observation

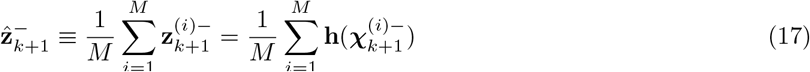

as well as the corresponding covariances

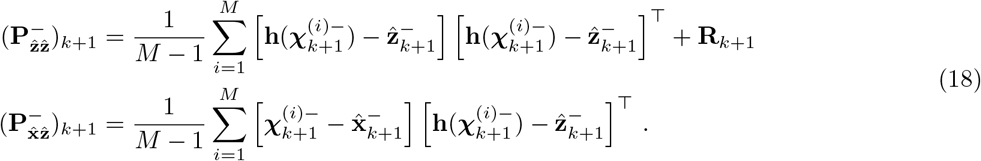

The Kalman gain is

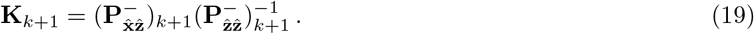

For a given observation **z**_*k*+1_, the state update of ensemble member *i* is

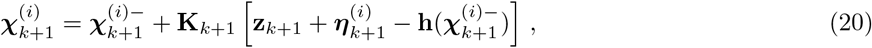

where 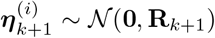 models Gaussian measurement noise. Finally, the updated state estimate and the corresponding covariance matrix are

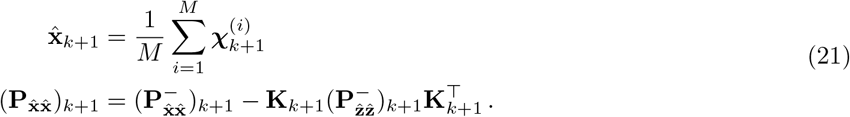

During each update step, we assign the entry in each ensemble member that corresponds to the logarithm of the drug-caused mortality rate, 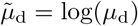, to be equal to the logarithm of the ratio of observed overdose fatalities and the ensemble mean of the population, while also accounting for the underlying noise.

### Overdose mortality data

Overdose fatalities extracted from the CDC WONDER database were identified using the International Classification of Diseases, Tenth Revision (ICD–10) cause-of-death codes X40–44 (unintentional), X60–64 (suicide), X85 (homicide), Y10–14 (undetermined intent), and all other drug-induced causes. National and county level data were extracted for the 1999–2020 period. The drug categories examined are poisoning by narcotics and psychodysleptics (hallucinogens) (T40) and by psychostimulants with abuse potential (T43.6). Specific subcategories analyzed within T40 are heroin (T40.1), natural and semisynthetic opioids (T40.2), and synthetic opioids other than methadone (T40.4). Deaths involving more than one drug type were included in each applicable category. Entries with an insufficient number of deaths were excluded.

## References

[1] Mattson, C. L. et al. Trends and geographic patterns in drug and synthetic opioid overdose deaths – United States, 2013–2019. Morbidity and Mortality Weekly Reports 70, 202–207 (2021).

[2] O’Donnell, J., Gladden, R. M., Mattson, C. L., Hunter, C. & Davis, N. L. Vital signs: Characteristics of drug overdose deaths involving opioids and stimulants - 24 states and the District of Columbia, January – June 2019. Morbidity and Mortality Weekly Reports 69, 1189–1197 (2020).

[3] Jones, C. M., Einstein, E. B. & Compton, W. M. Changes in synthetic opioid involvement in drug overdose deaths in the United States, 2010-2016. JAMA 319, 1819–1821 (2018).

[4] Armenian, P., Vo, K. T., Barr-Walker, J. & Lynch, K. L. Fentanyl, fentanyl analogs and novel synthetic opioids: A comprehensive review. Neuropharmacology 134, 121–132 (2018).

[5] Forman, R. F. & Block, L. G. The marketing of opioid medications without prescription over the internet. Journal of Public Policy and Marketing 25, 133–146 (2006).

[6] Mackey, T. K., Kalyanam, J., Katsuki, T. & Lanckriet, G. Twitter-based detection of illegal online sale of prescription opioid. American Journal of Public Health 107, 1910–1915 (2017).

[7] Lamy, F. R. et al. Listed for sale: Analyzing data on fentanyl, fentanyl analogs and other novel synthetic opioids on one cryptomarket. Drug and Alcohol Dependence 213, 108115 (2020).

[8] Duhart-Clarke, S. E., Kral, A. H. & Zibbell, J. E. Consuming illicit opioids during a drug overdose epidemic: Illicit fentanyls, drug discernment, and the radical transformation of the illicit opioid market. International Journal of Drug Policy 99, 103467 (2022).

[9] Kariisa, M., O’Donnell, J., Kumar, S., Mattson, C. L. & Goldberger, B. A. Illicitly manufactured fentanyl involved overdose deaths with detected xylazine – United States, January 2019 – June 2022. Morbidity and Mortality Weekly Reports 72, 721–727 (2023).

[10] Stein, E. M., Gennuso, K. P., Ugboaja, D. C. & Remington, P. L. The epidemic of despair among White Americans: Trends in the leading causes of premature death, 1999-2015. American Journal of Public Health 107, 1541–1547 (2017).

[11] Case, A. & Deaton, A. Deaths of despair and the future of capitalism (Princeton University Press, Princeton, NJ, 2020).

[12] Friedman, J. & Akre, S. COVID-19 and the drug overdose crisis: Uncovering the deadliest months in the United States, January – July 2020. American Journal of Public Health 111, 1284–1291 (2021).

[13] D’Orsogna, M. R., Böttcher, L. & Chou, T. Fentanyl-driven acceleration of racial, gender and geographical disparities in drug overdose deaths in the United States. PLOS Global Public Health 3, e0000769 (2023).

[14] Jalal, H. et al. Changing dynamics of the drug overdose epidemic in the United States from 1979 through 2016. Science 361, 6408 (2018).

[15] Peters, D. J., Monnat, S. M., Hochstetler, A. L. & Berg, M. T. The opioid hydra: Understanding overdose mortality epidemics and syndemics across the rural-urban continuum. Rural Sociology 85, 589–622 (2020).

[16] Powell, D., Shetty, K. D. & Peet, E. D. Trends in overdose deaths involving gabapentinoids and Z-drugs in the United States. Drug and Alcohol Dependence 249, 109952 (2023).

[17] Segel, J. E. & Winkelman, T. N. A. Persistence and pervasiveness: Early wave opioid overdose death rates associated with subsequent overdose death rates. Public Health Reports 136, 212–218 (2021).

[18] Blanco, C., Wall, M. M. & Olfson, M. Data needs and models for the opioid epidemic. Molecular Psychiatry 27, 787–792 (2022).

[19] Lim, T. Y. et al. Modeling the evolution of the US opioid crisis for national policy development. Proceedings of the National Academy of Sciences of the United States of America 119, e2115714119 (2022).

[20] Borquez, A. & Martin, N. K. Fatal overdose: Predicting to prevent. International Journal of Drug Policy 104, 103677 (2022).

[21] Monnat, S. M. Factors associated with county-level differences in U.S. drug-related mortality rates. American Journal of Preventive Medicine 54, 611–619 (2018).

[22] Rigg, K., Monnat, S. M. & Chavez, M. N. Opioid-related mortality in rural America: Geographic heterogeneity and intervention strategies. International Journal of Drug Policy 57, 119–129 (2018).

[23] Brownstein, J. S., Green, T. C., Cassidy, T. A. & Butler, S. F. Geographic information systems and pharmacoepidemiology: Using spatial cluster detection to monitor local patterns of prescription opioid abuse. Pharmacoepidemiological and Drug Safety 19, 627–637 (2010).

[24] Basak, A., Cadena, J., Marathe, A. & Vullikanti, A. Detection of spatiotemporal prescription opioid hot spots with network scan statistics: Multistate analysis. JMIR Public Health and Surveillance 5, e12110 (2019).

[25] Campo, D. S., Gussler, J. W., Sue, A., Skums, P. & Khudyakov, Y. Accurate spatiotemporal mapping of drug overdose deaths by machine learning of drug-related web-searches. PLOS One 15, e0243622 (2020).

[26] Marks, C. et al. Identifying counties at risk of high overdose mortality burden during the emerging fentanyl epidemic in the USA: A predictive statistical modelling study. Lancet Public Health 6, e720–e728 (2021).

[27] Marks, C. et al. Methodological approaches for the prediction of opioid use-related epidemics in the United States: A narrative review and cross-disciplinary call to action. Translational Research 234, 88–113 (2021).

[28] Sumetsky, N. et al. Predicting the future course of opioid overdose mortality: An example from two U.S. states. Epidemiology 61–69 (2021).

[29] Wagner, N. M. et al. Development and validation of a prediction model for opioid use disorder among youth. Drug and Alcohol Dependence 227, 108980 (2021).

[30] Stringfellow, E. J. et al. Reducing opioid use disorder and overdose deaths in the united states: A dynamic modeling analysis. Scientific Advances 8, eabm8147 (2022).

[31] Crassidis, J. L. & Junkins, J. L. Optimal estimation of dynamic systems (Chapman and Hall/CRC, Boca Raton, FL, 2004).

[32] Law, K., Stuart, A. & Zygalakis, K. Data assimilation: A mathematical introduction. Texts in Applied Mathematics (Springer, Cham, Switzerland, 2015).

[33] Böttcher, L., Chou, T. & D’Orsogna, M. R. Modeling and forecasting age-specific overdose mortality in the United States.European Physics Journal Special Topics (2023). URL 10.1140/epjs/s11734-023-00801-z.

[34] Bauer, P., Thorpe, A. & Brunet, G. The quiet revolution of numerical weather prediction. Nature 525, 47–55 (2015).

[35] Lillacci, G. & Khammash, M. Parameter estimation and model selection in computational biology. PLOS Computational Biology 6, e1000696 (2010).

[36] Schneider, T. et al. Epidemic management and control through risk-dependent individual contact interventions. PLOS Computational Biology 18, e1010171 (2022).

[37] Pei, S., Liljeros, F. & Shaman, J. Identifying asymptomatic spreaders of antimicrobial-resistant pathogens in hospital settings. Proceedings of the National Academy of Sciences of the United States of America 118, e2111190118 (2021).

[38] Li, R. et al. Substantial undocumented infection facilitates the rapid dissemination of novel coronavirus (SARS-CoV-2).Science 368, 489–493 (2020).

[39] Bomfim, R. et al. Predicting dengue outbreaks at neighbourhood level using human mobility in urban areas. Journal of the Royal Society Interface 17, 20200691 (2020).

[40] M’Kendrick, A. G. Applications of Mathematics to Medical Problems. Proceedings of the Edinburgh Mathematical Society 44, 98–130 (1925).

[41] Kermack, W. O. & McKendrick, A. G. Contributions to the mathematical theory of epidemics–I. 1927. Bulletin of Mathematical Biology 53, 33–55 (1991).

[42] Kermack, W. O. & McKendrick, A. G. Contributions to the mathematical theory of epidemics–II. The problem of endemicity. Bulletin of Mathematical Biology 53, 57–87 (1991).

[43] Kermack, W. O. & McKendrick, A. G. Contributions to the mathematical theory of epidemics–III. Further studies of the problem of endemicity. Bulletin of Mathematical Biology 53, 89–118 (1991).

[44] Diekmann, O., Othmer, H. G., Planqué, R. & Bootsma, M. C. J. The discrete-time Kermack–McKendrick model: A versatile and computationally attractive framework for modeling epidemics. Proceedings of the National Academy of Sciences of the United States of America 118, e2106332118 (2021).

[45] Xia, M., Greenman, C. & Chou, T. PDE models of adder mechanisms in cellular proliferation. SIAM Journal on Applied Mathematics 80, 1307–1335 (2020).

[46] Wang, Y., Dessalles, R. & Chou, T. Modeling the impact of birth control policies on China’s population and age: Effects of delayed births and minimum birth age constraints. Royal Society Open Science 9, 211619 (2022).

[47] Schenzle, D. An age-structured model of pre- and post-vaccination measles transmission. Mathematical Medicine and Biology 1, 169–191 (1984).

[48] Castillo-Chavez, C. & Feng, Z. Global stability of an age-structure model for TB and its applications to optimal vaccination strategies. Mathematical Biosciences 151, 135–154 (1998).

[49] Rong, L., Feng, Z. & Perelson, A. S. Mathematical analysis of age-structured HIV-1 dynamics with combination antiretroviral therapy. SIAM Journal of Applied Mathematics 63, 731–756 (2007).

[50] Böttcher, L., Xia, M. & Chou, T. Why case fatality ratios can be misleading: individual-and population-based mortality estimates and factors influencing them. Physical Biology 17, 065003 (2020).

[51] Chuang, Y. L., Chou, T. & D’Orsogna, M. R. Age-structured social interactions enhance radicalization. Journal of Mathematical Sociology 42, 128–151 (2018).

[52] Yang, J., Li, X. & Zhang, F. Global dynamics of a heroin epidemic model with age structure and nonlinear incidence. International Journal of Biomathematics 9, 1650033 (2016).

[53] Liu, L. & Liu, X. Mathematical analysis for an age-structured heroin epidemic model. Acta Applicandae Mathematicae 164, 193–217 (2019).

[54] Chekroun, A., Frioui, M. N., Kuniya, T. & Touaoula, T. M. Mathematical analysis of an age structured heroin-cocaine epidemic model. Discrete and Continuous Dynamical Systems - B 25, 4449–4477 (2020).

[55] Din, A. & Li, Y. Controlling heroin addiction via age-structured modeling. Advances in Difference Equations 521 (2020). URL 10.1186/s13662-020-02983-5.

[56] Duan, X., Cheng, H., Martcheva, M. & Yuan, S. Dynamics of an age structured heroin transmission model with imperfect vaccination. International Journal of Bifurcation and Chaos 31, 2150157 (2021).

[57] Khan, A., Zaman, G., Ullah, R. & Naveed, N. Optimal control strategies for a heroin epidemic model with age-dependent susceptibility and recovery-age. AIMS Mathematics 6, 1377–1394 (2021).

[58] Evensen, G. Sequential data assimilation with a nonlinear quasi-geostrophic model using Monte Carlo methods to forecast error statistics. Journal of Geophysical Research: Oceans 99, 10143–10162 (1994).

[59] Gompertz, B. On the nature of the function expressive of the law of human mortality, and on a new mode of determining the value of life contingencies. Philosophical Transactions of the Royal Society of London 115, 513–583 (1825).

[60] Makeham, W. M. On the law of mortality and the construction of annuity tables. Journal of the Institute of Actuaries 8, 301–310 (1860).

[61] Siler, W. A competing-risk model for animal mortality. Ecology 60, 750–757 (1979).

[62] Siler, W. Parameters of mortality in human populations with widely varying life spans. Statistics in Medicine 2, 373–380 (1983).

[63] Cohen, J. E., Bohk-Ewald, C. & Rau, R. Gompertz, Makeham, and Siler models explain Taylor’s law in human mortality data. Demographic Research 38, 773–841 (2018).

[64] Substance Abuse and Mental Health Services Administration, Results from the 2010 National Survey on Drug Use and Health: Summary of National Findings. NSDUH Series H-41, HHS Publication No. (SMA) 11-4658, Rockville, MD (2011).

[65] Mathers, B. M. et al. Mortality among people who inject drugs: a systematic review and meta-analysis. Bulletin of the World Health Organization 91, 102–123 (2013).

[66] Lindblad, R. et al. Mortality rates among substance use disorder participants in clinical trials: Pooled analysis of twenty-two clinical trials within the national drug abuse treatment clinical trials network. Journal of Substance Abuse Treatment 70, 73–80 (2016).

[67] Hser, Y. I. et al. High mortality among patients with opioid use disorder in a large healthcare system. Journal of Addiction Medicine 11, 315–319 (2017).

[68] Sordo, L. et al. Mortality risk during and after opioid substitution treatment: Systematic review and meta-analysis of cohort studies. BMJ 357, 357:j1550 (2017).

[69] King, C., Cook, R., Korthuis, P. T., Morris, C. & Englander, H. Causes of death in the 12 months after hospital discharge among patients with opioid use disorder. Journal of Addiction Medicine 16, 466–469 (2022).

[70] Bahji, A., Cheng, B., Gray, S. & Stuart, H. Mortality among people with opioid use disorder: A systematic review and meta-analysis. Journal of Addiction Medicine 14, e118–e132 (2020).

[71] Gini, C. Variabilità e mutabilità: Contributo allo studio delle distribuzioni e delle relazioni statistiche (C. Cuppini, Bologna, Italy, 1912).

[72] Xu, S., Böttcher, L. & Chou, T. Diversity in biology: definitions, quantification and models. Physical Biology 17, 031001 (2020).

[73] Courtwright, D. T. Dark Paradise: A History of Opiate Addiction in America (Harvard University Press, Cambridge, MA, 2001).

[74] Musto, D. F. The American disease: Origins of narcotic control (Oxford University Press, Oxford, UK, 1999), 3rd edn.

[75] Courtwright, D. T. A century of American narcotic policy. In Gerstein, D. R. & Harwood, H. J. (eds.) Treating drug problems, vol. 2 (National Academies Press, Washington, DC, 1992).

[76] Jones, C. M., Warner, M., Hedegaard, H. & Compton, W. Data quality considerations when using county-level opioid overdose death rates to inform policy and practice. Drug and Alcohol Dependence 204, 107549 (2019).

[77] Chou, T. & Greenman, C. D. A hierarchical kinetic theory of birth, death and fission in age-structured interacting populations. Journal of Statistical Physics 164, 49–76 (2016).

[78] Anderson, J. L. An ensemble adjustment Kalman filter for data assimilation. Monthly Weather Review 129, 2884–2903 (2001).

[79] Evensen, G. & Van Leeuwen, P. J. An ensemble Kalman smoother for nonlinear dynamics. Monthly Weather Review 128, 1852–1867 (2000).

[80] National Center for Health Statistics. NCHS Fact Sheet – June 2021: NCHS Data on Drug Overdose Deaths. Report (2021).

[81] Lipari, R. N. & Park-Lee, E. Mental Health Services Administration. Key substance use and mental health indicators in the United States: Results from the 2018 National Survey on Drug Use and Health (HHS Publication No. PEP19-5068, NSDUH Series H-54). Rockville, MD: Center for Behavioral Health Statistics and Quality. Substance Abuse and Mental Health Services Administration (2019).

[82] Brown, R. G. & Hwang, P. Y. C. Introduction to random signals and applied Kalman filtering: with MATLAB exercises and solutions (Wiley, Hoboken, NJ, 1997).

[83] Labbe, R. Kalman and Bayesian Filters in Python, https://github.com/rlabbe/Kalman-and-Bayesian-Filters-in-Python/blob/master/Appendix-E-Ensemble-Kalman-Filters.ipynb (2022).

[84] Brown, R. G. & Hwang, P. Y. C. Introduction to random signals and applied Kalman filtering: with MATLAB exercises and solutions (Wiley, Hoboken, NJ, 2012).

